# Is the Patient State Index Recorded with the Sedline Sedation Monitor Correlated with the Duration of Emergence After Pediatric Surgery: An Exploratory, Single-center, Blinded, Prospective Cohort Study

**DOI:** 10.1101/2023.02.27.23286232

**Authors:** Ludmil Mitrev, Stuart Pasch, Ian Brotman, Bharathi Gourkanti, Fatima Habib, Michael Schwartz, Noud van Helmond

## Abstract

**Introduction:** Following surgery with general anesthesia, some children experience a prolonged emergence. We designed a prospective observational study in children undergoing general anesthesia who were monitored with the SedLine^®^ Sedation Monitoring system (Masimo Corporation, Irvine, CA) to explore the hypothesis that the Patient State Index (PSI) obtained with this monitor could be inversely correlated with the duration of emergence after anesthesia.

**Materials and methods:** Prospective, observational single center study in a tertiary academic center in the United States. Fifty-six children between the ages of 1 and 12 years scheduled to undergo non-emergent surgery with general inhalational anesthesia were enrolled. Demographic and intraoperative characteristics were recorded. All caregivers were blinded to the PSI. Correlations were derived between PSI, duration of emergence, post-anesthesia care unit length of stay (LOS), and hospital LOS. PSI was analyzed in categories of <25, 25-50, and >50 both as absolute time spent in each category, and as the fraction of time compared to the length of the anesthetic. The development of emergence delirium (ED) was recorded as a secondary outcome variable.

**Results:** The correlation coefficients between the PSI categories and the outcomes were weak (<0.3). Only two of the correlation coefficients reached statistical significance at p=0.05: fraction and absolute time spent in PSI category > 50 and PACU length of stay, indicating that longer periods of PSI > 50 during the anesthetic were associated with longer PACU LOS. Three patients (5%) developed ED.

**Conclusion:** PSI measured with the SedLine monitor was not significantly correlated with the duration of emergence. There was a weak positive correlation between intraoperative time spent with PSI readings >50 and PACU LOS. Our sample did not have a high-enough event rate of ED to make statistical inferences about a correlation between PSI, ED and duration of emergence.

## Introduction

After surgery with general anesthesia, some children experience a prolonged emergence. Risk factors for prolonged pediatric emergence are poorly researched. We designed a prospective observational study to determine whether the Patient State Index (PSI) obtained with the SedLine^®^ Sedation Monitoring system (Masimo Corporation, Irvine, CA) correlated with prolonged emergence from general anesthesia in this population. The SedLine monitor uses four EEG electrodes to record frontal and pre-frontal EEG signals, which it then processes to derive a PSI parameter: a validated measure of the effect (depth) of anesthesia. PSI is a whole number ranging from 0 (deep anesthesia) and 100 (awake state) calculated by an algorithm that takes into account the power of the EEG frequency bands, the phase information from anteroposterior synchronization and the bilateral coherence of the brain, and the inhibition of frontal cortical regions. ^1−3^ The suggested range for general anesthesia is 25-50. ^1^ The exact calculation of PSI is based on proprietary, empirical algorithms. The basic principles of the algorithms are published. ^4^ The SedLine monitor also visualizes the power of the four main EEG frequency bands (beta, alpha, theta and delta) on screen using a color-coded density spectral array. ^5^

Our study hypothesis was that lower PSI values for longer periods of time during anesthesia would be correlated with longer emergence times. If this were found to be the case, the PSI number could potentially prove a useful parameter for predicting delayed pediatric emergence. This might allow for more precise titration of the anesthesia and shorter exposure of the child to the operating room environment.

## Materials and Methods

This was a single institution, prospective, observational, non-interventional study. The study was approved by our institutional review board (IRB # 19-166ES). From June 1^st^, 2020 to July 30^th^, 2021, all pediatric patients between the ages of 1 and 12 years who were scheduled to undergo surgery with general inhalational anesthesia were approached for enrollment. Written informed consent was provided by parents or legal guardians and assent was obtained from minors old enough to provide it. Only screened cases who provided written informed consent were enrolled.

Besides age, the inclusion criteria consisted of American Society of Anesthesiologists (ASA) physical status I or II. The exclusion criteria were emergency surgery, chronic treatment with central nervous system drugs (antiepileptics, antidepressants, sedatives), cerebral palsy, seizure disorder or status epilepticus, Down syndrome, and anticipated case duration less than 40 minutes.

On arrival to the operating room, the patients were attached to a SedLine monitor to assess PSI with the electrodes applied as recommended by the manufacturer. The SedLine system is comprised of four components: Root (monitor box with screen), module, patient cable, and sensors. The information is displayed on the Root monitor. This includes electrode status, EEG waveforms, PSI, electromyograph (EMG), artifacts (ARTF), and suppression ratio (SR). The EEG display reflects electrical activity of the frontal and pre-frontal cortex of the brain. Electromyography (EMG) is a measure of detected muscle activity, such as grimacing or jaw clenching. EMG is represented by a numeric value that ranges from 0 to 100%. When an EMG numeric value is not available, the value displays dashes (--). The Suppression Ratio (SR) is a measure of how much the electrical activity of the frontal and pre-frontal cortex of the brain is suppressed as a percentage of time. Artifact (ARTF) is a measure of how much physiological (non-brain related) and environmental noise is detected by the system. ^5^

The SedLine sensor is comprised of six gelled electrodes, including four active channels (R1, R2, L1, L2), one reference channel (CT), and one ground channel (CB). The sensor is a single-use, non-sterile product that does not contain natural rubber latex. ^5^

In addition to the SedLine monitor, standard anesthesia monitors were applied. The SedLine screen was covered or turned away from the anesthesia provider, surgeon and patient, who were blinded to the monitor output. PSI over time was downloaded from the monitor at the end of each case by research staff and transferred to a Microsoft Excel spreadsheet for pre-analysis. Demographic data and intraoperative drug administration data was also recorded.

The primary end point was the duration of anesthesia emergence (in minutes) with secondary end points including: incidence of emergence delirium (ED), incidence of post-operative nausea and vomiting, post-anesthesia care unit (PACU) length of stay, and hospital length of stay. ED was assessed clinically by the attending anesthesiologist via direct observation of the patient at the end of surgery and through the early post-operative period.

### Statistical analysis

To detect a correlation of |.60| or greater between the PSI and the duration of emergence, a minimum sample size of 57 was required. The sample size was calculated based on a null correlation of 0.25, 90% power and a p-value of 0.05. In anticipation of data loss or subject dropout, we received approval for, and enrolled, 60 subjects into the study.

Demographic and intraoperative characteristics were expressed as means with standard deviations (SD) or proportions (%). The PSI was divided into range categories of <25, 25-50, and >50. The duration spent in each category was calculated and added up to yield an absolute number in minutes, as well as expressed as a fraction of the total duration of the anesthetic. Spearman correlations were calculated between these predictor variables and the duration of emergence, hospital length of stay and PACU length of stay. Sigmaplot statistical software was used (Inpixon - systatsoftware.com, Palo Alto, CA).

## Results

The baseline characteristics of the cohort are summarized in Table 1. Table 2 summarizes the perioperative characteristics of the sample. The mean emergence duration was 11 minutes. Three subjects (5%) had signs and symptoms indicative of ED. The mean PACU length of stay was 137 minutes.

**Table 1.** Baseline cohort characteristics.

|  | Analysis cohort |
| --- | --- |
| Sample size | 56 |
| Age in years, mean ± SD | 6.3 ± 3.2 |
| Male sex, n (%) | 38 (68) |
| Body Mass Index in kg/m<sup>2</sup>, mean ± SD | 18 ± 4.5 |
| Type of surgery, n (%) |  |
| Abdominal surgery | 5 (9) |
| General surgery of soft tissue (e.g. lymph node excision) | 5 (9) |
| Hernia surgery | 24 (43) |
| Oromaxillary/Otolaryngology surgery | 9 (16) |
| Orthopedic surgery | 9 (16) |
| Urologic surgery | 4 (7) |

SD = standard deviation

**Table 2.** Perioperative events and characteristics of the study cohort.

|  | Analysis cohort |
| --- | --- |
| Duration of anesthesia in minutes, mean ± SD | 77 ± 42 |
| Emergence time in minutes, mean ± SD | 11 ± 7 |
| Postoperative nausea and vomiting, n (%) | 3 (5) |
| Emergence delirium, n (%) | 3 (5) |
| PACU length of stay in minutes, mean ± SD | 137 ± 47 |
| Hospital length of stay in days, median (range) | 0 (0 – 3) |

SD = standard deviation; PACU = post-anesthesia care unit

Figure 1 is a box plot showing the proportion of time the subjects in the cohort spent in each PSI range category. The three subjects who experienced pediatric ED are depicted by solid circles. Outliers are depicted as open circles. The distribution of the subjects experiencing ED showed that they were not clustered in a particular fashion.

**Figure 1.**
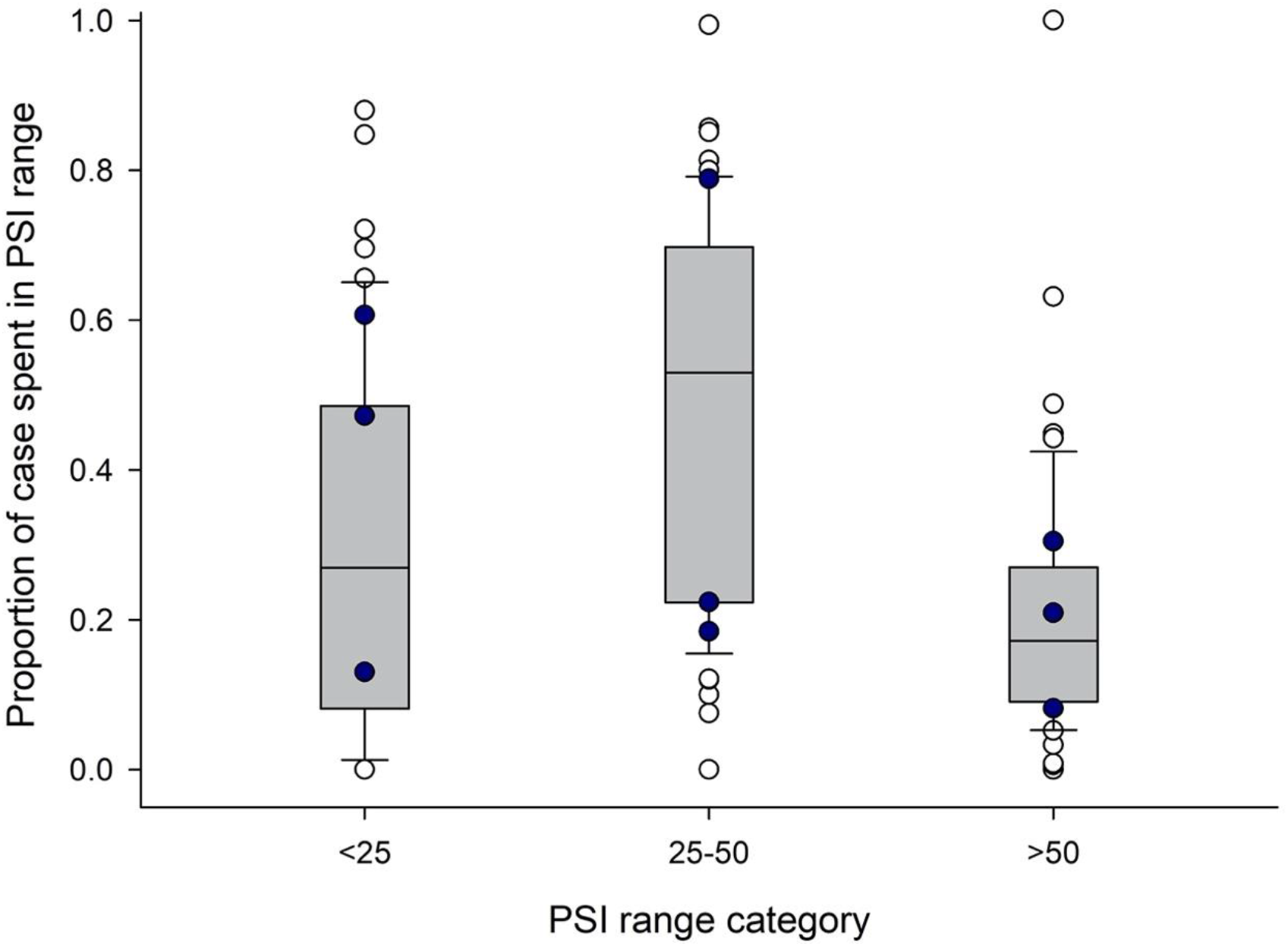
Box plot of proportion of time spent in each PSI category. Solid circles: emergence delirium; Open circles: outliers; PSI = patient state index

The Spearman correlation coefficients of the PSI categories and the duration of emergence are shown in Table 3. All correlation coefficients were weak (<0.3). Only two of the correlation coefficients reached statistical significance at p=0.05: fraction and absolute time spent in PSI category > 50 and PACU length of stay. This indicated that longer periods of PSI>50 during the anesthetic were associated with longer PACU LOS.

**Table 3.**
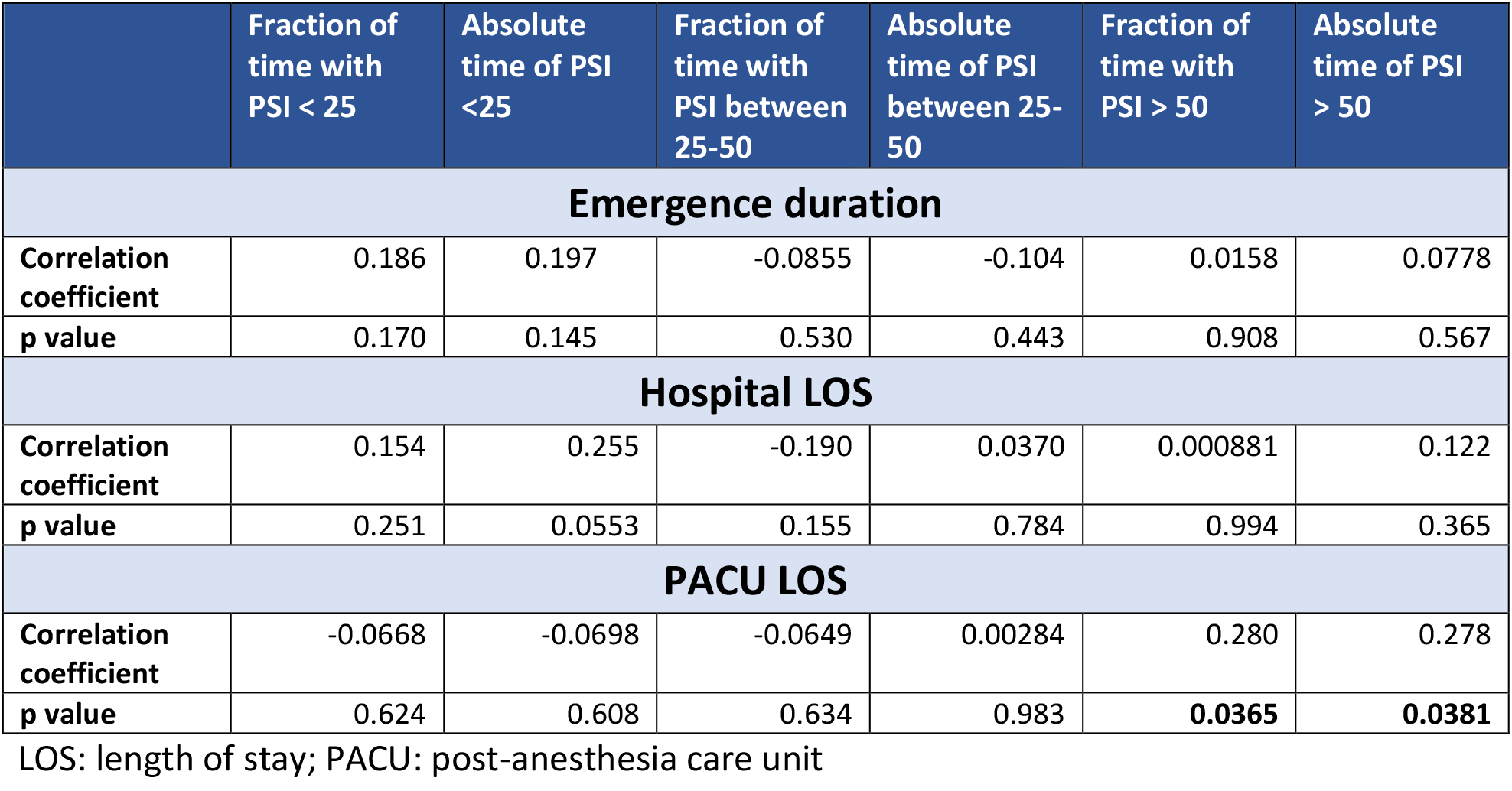
Spearman correlation between PSI parameters and emergence time, hospital and PACU LOS.

## Discussion

The PSI was not significantly correlated with the duration of emergence after pediatric anesthesia, hospital LOS or PACU LOS, except for a mild positive correlation of PSI > 50 with PACU LOS. The Spearman correlation coefficients were 0.280 and 0.278 for the fraction of time spent with a PSI >50 and the absolute time with a PSI >50, respectively. This correlation did not extend to PSI values of <25 or 25-50.

Anesthesiologists have been using intraoperative neuromonitoring to assist in anesthetic titration for decades. More recently, machine algorithms have been introduced to reduce the skill burden on interpretation of raw EEG. The peri-anesthetic use of EEG has been well studied in the adult population. ^6−9^ However, there is a paucity of data in the pediatric population.

Modern multi-modal anesthesia combining anesthetics, opioids and other adjuvants carries the potential for wide-ranging emergence times. We postulated that deeper anesthesia might correlate with longer emergence in this patient population, but our data did not bear this out.

ED is an important condition that sometimes occurs after surgery. Its clinical features include inconsolable crying, non-purposeful movements, disorientation, lack of eye contact, aggressiveness or irritability, and non-responsiveness, while its major predisposing factors are anxiety prior to surgery, preschool age, and sevoflurane or desflurane use. ED can lead to patient harm such as untimely removal of intravenous catheters, and to delays in PACU stay. ^10^ Our goal with recording the incidence of ED was to examine its correlation with the duration of emergence and PSI, assuming that enough cases of ED occurred in our sample. The incidence of ED across all age groups is between 5.3% and 50%, but in children, it has been reported at 12-13%.^11^ Our study recorded 3 instances of delirium (5%), in the lower range of published data. We found no specific trend for ED and PSI category. A larger sample would be required to make inferences about the correlation between ED, PSI and the duration of emergence. Other authors have reported that rapid emergence from anesthesia is a risk factor for ED. ^12,13^ In addition, at least one randomized controlled trial attempted to elucidate whether depth of anesthesia was associated with ED, but the results were inconclusive. ^14^ A future study, adequately powered, would need to be carried out to ascertain whether controlling depth of anesthesia via the PSI can play a role in reducing the incidence of ED.

## Conclusions

PSI measured with the SedLine monitor was not significantly correlated with the duration of emergence in a blinded single-center prospective cohort of 56 pediatric patients undergoing non-emergent surgery with general anesthesia. There was a weak positive correlation between intraoperative time spent with PSI readings >50 and PACU LOS. Our sample did not have a high-enough event rate of ED to make statistical inferences about a correlation between PSI, ED and duration of emergence.

## Data Availability

All data produced in the present work are contained in the manuscript.

